# Agreement between cystatin-C and creatinine based estimated glomerular filtration rate among Ethiopian children

**DOI:** 10.64898/2026.03.05.26347688

**Authors:** Beakal Zinab, Rahma Ali, Bikila S. Megersa, Tsinuel Girma, Henrik Friis, Mubarek Abera, Tefera Belachew, Mette Frahm Olsen, Suzanne Filteau, Jonathan CK. Wells, Rasmus Wibaek, Daniel Yilma, Dorothea Nitsch

## Abstract

**Introduction:** Glomerular filtration rate (GFR) is invasive to measure. Therefore, in clinical care, estimated GFR is derived from serum levels of endogenous filtration markers such as creatinine and cystatin C. Multiple studies from high income countries showed differences between estimated glomerular filtration rate based on cystatin C (eGFRcys) and creatinine (eGFRcr). This study aimed to assess the agreement between eGFRcys and eGFRcr in Ethiopian children and identify factors influencing higher eGFRcys and eGFRcr.

**Method:** We studied 350 Ethiopian children who were part of the iABC birth cohort study. At the recent follow-up (average age 10 years), serum cystatin C and creatinine were measured. Formulas by Berg (2015) and Hoste (2014) were used to estimate eGFRcys and eGFRcr, respectively, and Bland–Altman plots assessed their agreement. The difference in eGFR (eGFRdiff) was calculated and categorized as less than-15 mL/min/1.73 m² (higher eGFRcr), between-15 and <15 mL/min/1.73 m² (concordant), and greater than or equal to 15 mL/min/1.73 m² (higher eGFRcys). Multinomial logistic regression was used to identify factors associated with higher eGFRcr and higher eGFRcys.

**Result:** Estimated glomerular filtration rate (eGFR) showed significant variation based on the estimation formula used. When using formulas by Berg (2015) and Hoste (2014), the median (IQR) eGFRcys and eGFRcr were 99.4 (90.0; 114.1), and 123.2 (110.3; 143.8) mL/min/1.73 m^2^, respectively. Overall, we observed a poor agreement between eGFRcys and eGFRcr, with only 94 (27.6%) children having concordant results compared to 220 (64.7%) with higher eGFRcr and 26 (7.6%) with higher eGFRcys. If the eGFRcys results are considered reliable, 27.5% of the children had eGFR below 90 mL/min/1.73 m².

**Conclusion:** There was very marked variation in the distributions of estimated eGFRs depending on which formulas for children were used. Agreement between eGFR estimated using cystatin C and creatinine was poor among Ethiopian children. Relative to eGFRcys, kidney function may be overestimated by creatinine-based equation as up to 30ml/min in Ethiopia. Ideally, a validation study with GFR measured by gold standard methods (Inlulin clearance) among children is required. However, because of its invasive nature and financial concerns, Iohexol clearance studies are recommended.

## Introduction

Chronic kidney disease (CKD) is increasingly recognized as a significant health problem, not only in high-income countries but also in low-income regions like sub-Saharan Africa (SSA) (1). To date it is unknown why there is such a high burden of CKD in SSA. One hypothesis is that kidney problems start in childhood. Abnormal urinary markers of kidney disease are very common among children living in sub-Saharan Africa (1,2). Prenatal kidney injury, childhood malnutrition, and acute kidney disease (AKD) that associated with the prevalent childhood diarrhea may contribute to the development of adult chronic kidney disease (CKD). However, longitudinal studies are needed to confirm that such changes track to adulthood CKD (3,4).

To understand how kidney function in children relates to kidney function in adults, accurate determination of glomerular filtration rate (GFR) is needed, and this would also help clinical identification of pediatric kidney diseases (5). While precise measurement of GFR can be achieved through exogenous marker clearance studies, their invasive and time-consuming nature limits their widespread use. Consequently, GFR is typically estimated (eGFR) from serum concentrations of endogenous markers such as creatinine or cystatin C in routine clinical settings (6–10). To this end, multiple eGFR equations have been developed for both adult and pediatric populations (7,10,11) and eGFR is usually normalized to a body surface area of 1.73 m^2^ (12). Studies indicated cystatin C may be a more sensitive marker of changes in GFR than serum creatinine (13,14).

Discrepancies between eGFR calculated using cystatin C (eGFRcys) and creatinine (eGFRcr), termed eGFRdiff, can be observed when both markers are measured simultaneously in the same individual (7,15). Such differences between eGFRcys and eGFRcr are likely attributable to factors extrinsic to renal function that differentially influence cystatin C or creatinine serum concentrations (15). These factors may reflect muscle or fat mass, activity level, or chronic inflammation (10,16,17).

In adults in high income settings, lower eGFRcys relative to eGFRcr has been associated with increased risk of frailty, heart failure, hospitalizations, cardiovascular disease events, kidney failure, and mortality (18–20). In children in high income settings, eGFRcys is the preferred kidney function marker because it is thought not to be affected by growth or muscle mass, unlike eGFRcr, which needs to take these factors into account. A recent gold-standard validation study from SSA in adults found that eGFRcr misses those with reduced kidney function, in contrast to eGFRcys (21,22).

Although there are multiple recommended estimation formulas available for children, which GFR estimation formula should be used in children from SSA is unclear, leading to uncertainty in clinical decision making (15,23,24). The current study aimed to i) describe the distributions of different recommended pediatric eGFR formulas in normal Ethiopian children, ii) assess the level of agreement between eGFRcys and eGFRcr and iii) identify factors associated with any discrepancy between eGFR estimated with cystatin C and creatinine.

## Methods

### Study setting and participants

This study included children from the 10-year follow-up of the infant anthropometry and body composition (iABC) birth cohort from Jimma Town, Ethiopia. The cohort has been described elsewhere (25,26) and had a total of 12 prior study visits. The 10-year follow-up visits were conducted between June 2019 and December 2020 among 350 children aged 7-12 years. Families were contacted using previously provided phone numbers or by visiting their last known addresses. Parents were invited to bring their children for the study after receiving detailed information.

### Blood sample collection and analysis

Families were instructed to ensure their children fasted for eight hours prior to the visit in the morning. Upon arrival, laboratory technicians verified the child’s fasting status before collecting a 4 ml blood sample. The study nurses provided the children with a small meal immediately after sample collection. The samples were stored in K2-EDTA tubes in the lab fridge for no more than 4 hours, then centrifuged and stored at-80°C until further analysis. Serum cystatin C was determined using an enhanced immune turbidimetric assay and serum creatinine level using Jaffe with an alkaline picrate assay method, both on a Cobas c 702 analyzer (Roche Diagnostic, Germany).

### Estimation of glomerular filtration rate (GFR)

As there is no validated pediatric eGFR formula for low-income countries, multiple eGFR estimation formulas were initially assessed before selecting the most suitable one (Supplemental Table 1). The Berg 2015 and Hoste 2014 formulas were ultimately selected due to their relative closeness in the iABC population and superior P_30_ accuracy in past validation studies (27). P_30_ is the percentage of individuals with eGFR within +30% of measured GFR (28). Therefore, in this study we estimated GFR using the following formulas:

Creatinine-based eGFR: 107.3/(sCr/Q) (formula by Hoste et al., 2014); Q= 0.0270*age in years+0.2329 (29). Cystatin C-based eGFR: 91 x sCys (mg/L) ^-1.213^ (formula by Berg et al., 2015) (30)

### Level of discrepancy between eGFRcys and eGFRcr

The eGFRdiff, defined as eGFRcys minus eGFRcr, was categorized into 3 levels: higher eGFRcr (eGFRdiff < −15 mL/min/1.73 m^2^), concordant eGFRcys and eGFRcr (eGFRdiff-15 to < 15 mL/min/1.73 m^2^), and higher eGFRcys (eGFRdiff≥ 15 mL/min/1.73m^2^). These eGFRdiff cutoffs were chosen because 15 mL/min/1.73 m^2^: i) corresponds to approximately 1-standard deviation of eGFRdiff, ii) represents clinical significance, and iii) has been used in prior similar studies (15,20).

### Predictor variables

Based on evidence from prior literature, covariates potentially associated with eGFRdiff were identified. Candidate variables were categorized under three domains: biological (age and sex), anthropometric (height, fat mass and fat-free mass at the 10-year follow-up) and clinical (C-reactive protein) characteristics. Weight was measured to the nearest 0.1 kg using an electronic UNICEF scale (Seca). Standing height was measured to the nearest 0.1 cm using a Seca 213 portable stadiometer (Seca). Body composition was measured using air displacement plethysmograph (BOD POD; COSMED, Rome, Italy). The BOD POD has high accuracy and precision and is feasible and safe for assessing body composition in infants and children (31).

## Statistical analyses

Data were double entered into Epi Data version 4.4.2.0 (Odense, Denmark). Data cleaning and further analysis were done using Stata version 15 (Stata Corp LLC., College Station, Texas, USA). The descriptive data were presented as mean (standard deviation [SD]) and median (interquartile range [IQR]) for normally and non-normally distributed data, respectively, and count (percentage) for categorical variables. Because there were very few children classified as having eGFR<90ml/min using Hoste 2014 creatinine-based equations (n=7), we described data using an eGFR cutoff of 100ml/min for both eGFRcr and eGFRcys. Bland-Altman plots were used to evaluate the bias and agreement between cystatin C and creatinine-based eGFR estimations. The mean bias percentage was calculated by dividing the difference between eGFRcys and eGFRcr by their average and then multiplying by 100%. Multinomial logistic regression analysis was used to identify factors associated with eGFRdiff. The concordant category was used as a reference group to compare higher eGFRcys or eGFRcr categories. Two multinomial models were fitted: Model 1 included age and sex and Model 2 additionally adjusted for fat mass, fat-free-mass and C-reactive protein at age 10 years.

Sensitivity analyses were carried out for Schwartz (2009) formulas, as these are currently used in clinical care in Ethiopia, as well as exploring the influence of very high eGFR estimates on the results.

### Ethics

Ethical approval for the study was granted by Jimma University Ethical Review Board of the College of Public Health and Medical Sciences (reference IHRPHD/333/18). Parents/guardians signed consent after getting detailed information regarding the study was provided. Any abnormal findings detected during clinical and laboratory evaluations were communicated to families of children and they were linked to Jimma University Medical Center for further evaluation.

## Results

### Characteristics of mothers and children

A total of 644 mother-infant pairs were initially involved in the study. However, 10 preterm babies and 63 children who did not live in Jimma were not included in follow-up visits. Of the 571 eligible for follow-up studies, 350 (61.3%) participated in the 10-year follow-up to assess kidney outcomes. Of these 350 children, serum cystatin C and creatinine measurements were taken for 341 and 340, respectively (Supplemental Figure 1). Of those mothers, 39% had attended secondary school, and 22.3% were in the poorest wealth index category. Of the 350 children, 51.4% were male, the mean (±SD) age was 9.8 ± 1.0 y, and 48.6% were firstborn. Median serum creatinine and cystatin C in this cohort were 0.43 (IQR 0.38-0.48) mg/dL, and 0.93 (IQR 0.83-1.01) mg/L, respectively.

Comparing children with higher eGFR readings to those with lower eGFR readings (using 100ml/min/1.73m^2^ as a cut-off value) in Table 1, there was a slight shift to more boys and children born to less wealthy mothers being in the higher eGFR groups.

**Table 1:** Maternal and child characteristics of children with higher/lower cystatin C and creatinine.

|  | eGFRcys (Berg) |  | eGFRcr (Hoste) |  | Missing,<br>n |
| --- | --- | --- | --- | --- | --- |
|  | Lower (N=181)<br>≤100ml/min/1.72m² | Higher (N=160)<br>>100ml/min/1.72m² | Lower (N=31)<br>≤100ml/min/1.72m² | Higher (N=309)<br>>100ml/min/1.72m² |  |
| Maternal characteristics |  |  |  |  |  |
| Educational status, n (%) |  |  |  |  | 4 |
| No school | 9 (5.0) | 11 (7.0) | 1 (3.3) | 19 (6.2) |  |
| Primary school | 60 (33.1) | 44 (28.2) | 3 (10.0) | 101 (33.0) |  |
| Secondary school | 67 (37.0) | 67 (43.0) | 16 (53.3) | 118 (38.6) |  |
| Higher education | 45 (24.9) | 34 (21.8) | 10 (33.3) | 68 (22.2) |  |
| Wealth index, n (%) |  |  |  |  | 0 |
| Poorest | 30 (16.6) | 47 (29.4) | 4 (13.0) | 73 (23.6) |  |
| Poorer | 30 (16.6) | 31 (19.4) | 1 (3.2) | 60 (19.4) |  |
| Middle | 47 (26.0) | 21 (13.1) | 7 (22.6) | 61 (19.7) |  |
| Rich | 36 (19.8) | 35 (21.9) | 7 (22.6) | 64 (20.7) |  |
| Richest | 38 (21.0) | 26 (16.2) | 12 (38.8) | 51 (16.6) |  |

| **Child characteristics at birth** |  |  |  |  |  |
| --- | --- | --- | --- | --- | --- |
| BW (kg), mean (SD) | 3.0 (0.4) | 3.0 (0.4) | 3.1 (0.5) | 3.0 (0.4) | 4 |
| Gestation (wks), mean (SD) | 39.0 (1.0) | 39 (0.9) | 38.8 (0.8) | 39.0 (0.9) | 0 |
| Birth order, n(%) |  |  |  |  | 2 |
| First born | 89 (49.4) | 75 (47.2) | 15 (50.0) | 148 (48.0) |  |
| Second born | 44 (24.4) | 48 (30.2) | 8 (26.7) | 84 (27.3) |  |
| Third and above | 47 (26.1) | 36 (22.6) | 7 (23.3) | 76 (24.7) |  |
| **Child characteristics at 10-year follow-up** |  |  |  |  |  |
| Age (yrs), mean (SD) | 9.9 (1.0) | 9.6 (0.9) | 9.3 (1.0) | 9.8 (1.0) | 0 |
| Sex, n(%) |  |  |  |  | 0 |
| Male | 90 (49.7) | 87 (54.4) | 14 (45.2) | 163 (52.7) |  |
| Female | 91 (50.3) | 73 (45.6) | 17 (54.8) | 146 (47.3) |  |
| Height (cm), mean (SD) | 133.8 (8.1) | 130.3 (6.6) | 132.0 (8.7) | 132.1 (7.6) | 0 |
| Weight (kg) mean, (SD) | 28.5 (6.9) | 25.8 (4.4) | 26.6 (6.9) | 27.3 (5.9) | 0 |
| FM (kg), mean (SD) | 6.0 (4.2) | 5.0 (2.4) | 5.2 (4.1) | 5.6 (3.5) | 2 |
| FFM (kg), mean (SD) | 22.4 (3.5) | 20.9 (2.9) | 21.5 (3.6) | 21.7 (3.3) | 2 |

Data are mean (SD) for continuous and count (%) for categorical variables. Unless otherwise specified, column percentages are shown in brackets. Abbreviations: FM, fat mass, FFM, fat-free mass, BW, birth weight.

### Distribution of eGFR estimated using multiple cystatin C and creatinine-based formulas

There was wide discrepancy in the distribution of eGFR depending on the estimation formula used for the same children across a wide range (Figure 1). Both 2012 Schwartz equations put the entirety of this normal cohort of Ethiopian children into a reduced kidney function range, whilst other equations suggested that more of these children have normal kidney function. Creatinine-based Schwartz 2009 (used in clinical care in Ethiopia) and Hoste 2014 formulas produced comparable GFR distributions, as did cystatin C-based Berg 2015 and Grubb 2014 formulas (Figure 1).

**Figure 1:**
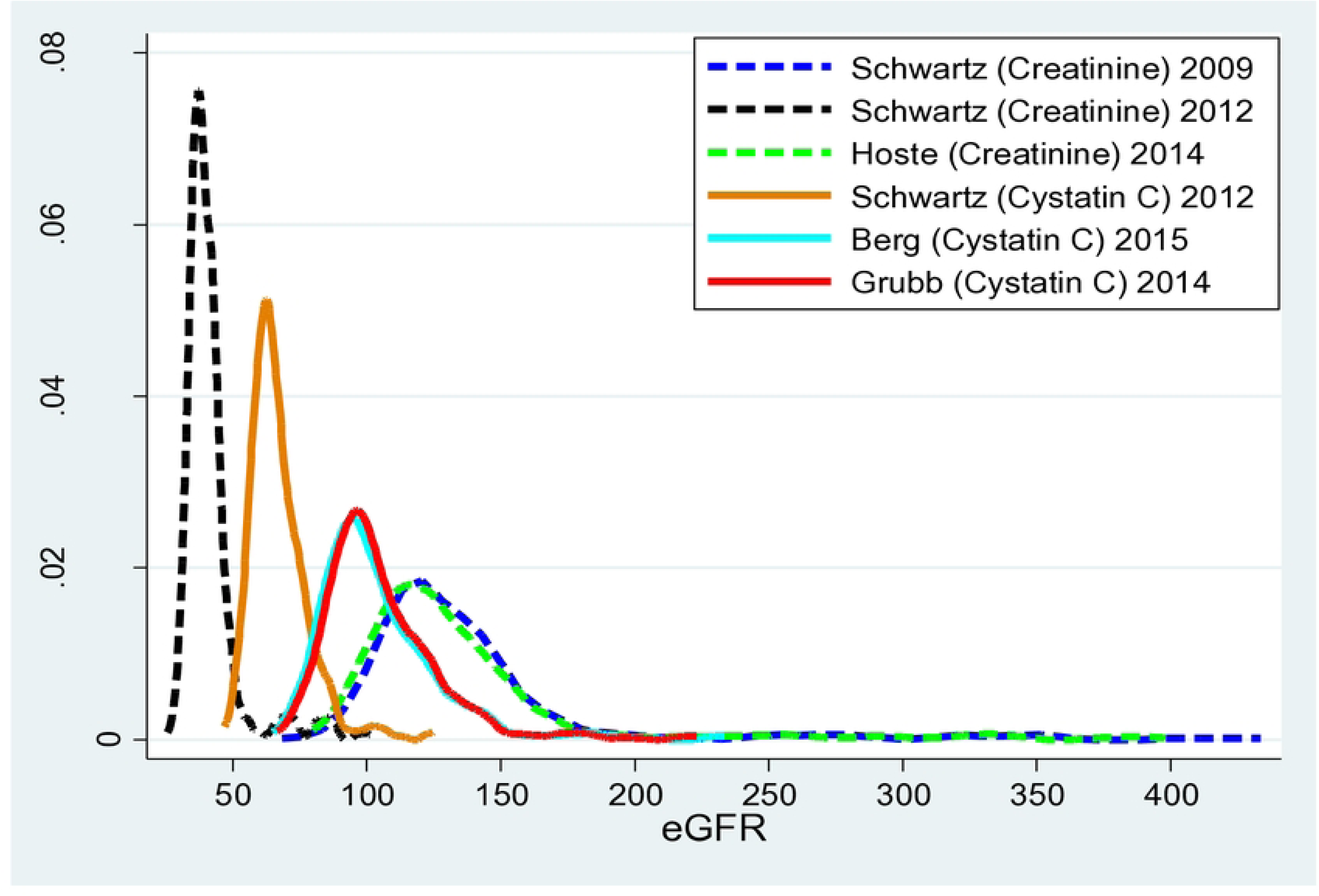
Comparison of distributions of GFR estimated using different formulas Kernel Density Distribution plot. GFR glo1nerular filtration rate. Schwartz (creatinine) 2009 (32) and 2012=Schwartz GFR esti1nation creatinine equation adjusted for height (33). Hoste (creatinine) 2014=creatinine based GFR estimation equation adjusted for age (29). Schwartz (cystatin C) 2012=GFR estin1ation cystatin C equation (33). Berg (cystatin C) 2015=GFR estimation cystatin C equation (30). Grubb (cystat.in C) 2014=GFR estinlation cystatin C equation (34)

The median (IQR) of eGFR estimated using cystatin C (Berg 2015) and creatinine (Hoste 2014) based formula were 99.4 (90.0-114.1) mL/min/1.73 m^2^, and 123.2 (110.3-143.8) mL/min/1.73 m^2^, respectively. If the Berg 2015 equation is considered as being accurate, 94 (27.5%) of the children in this study have an estimated GFR below 90 mL/min/1.73m². Children with an eGFR below 90 mL/min/1.73 m², estimated using the cystatin C-based Berg 2015 equation, were slightly older and heavier than those with higher eGFR (Supplemental Table 2),

### Agreement between eGFR estimated with cystatin C (Berg 2014) and creatinine (Hoste 2014) based formulas

The median (IQR) eGFR difference between cystatin C based and creatinine based eGFR was - 25.2 (-41.3;-9.3) ml/min/1.73 m^2^. The overall mean bias percentage was-23.7% with limits of agreement (LOA) estimated to be-79.8 and 33.8 % (Figure 2-A). The overall average bias was - 30.9 ml/min/1.73m^2^ with LOA of-127.7 and 65.9 ml/min/1.73m^2^. Comparatively better agreement was observed when mean GFR < 150ml/min/1.73m^2^, whereas there was variability in both directions at >150 ml/min/1.73m^2^ (Figure 2-B).

**Figure 2.**
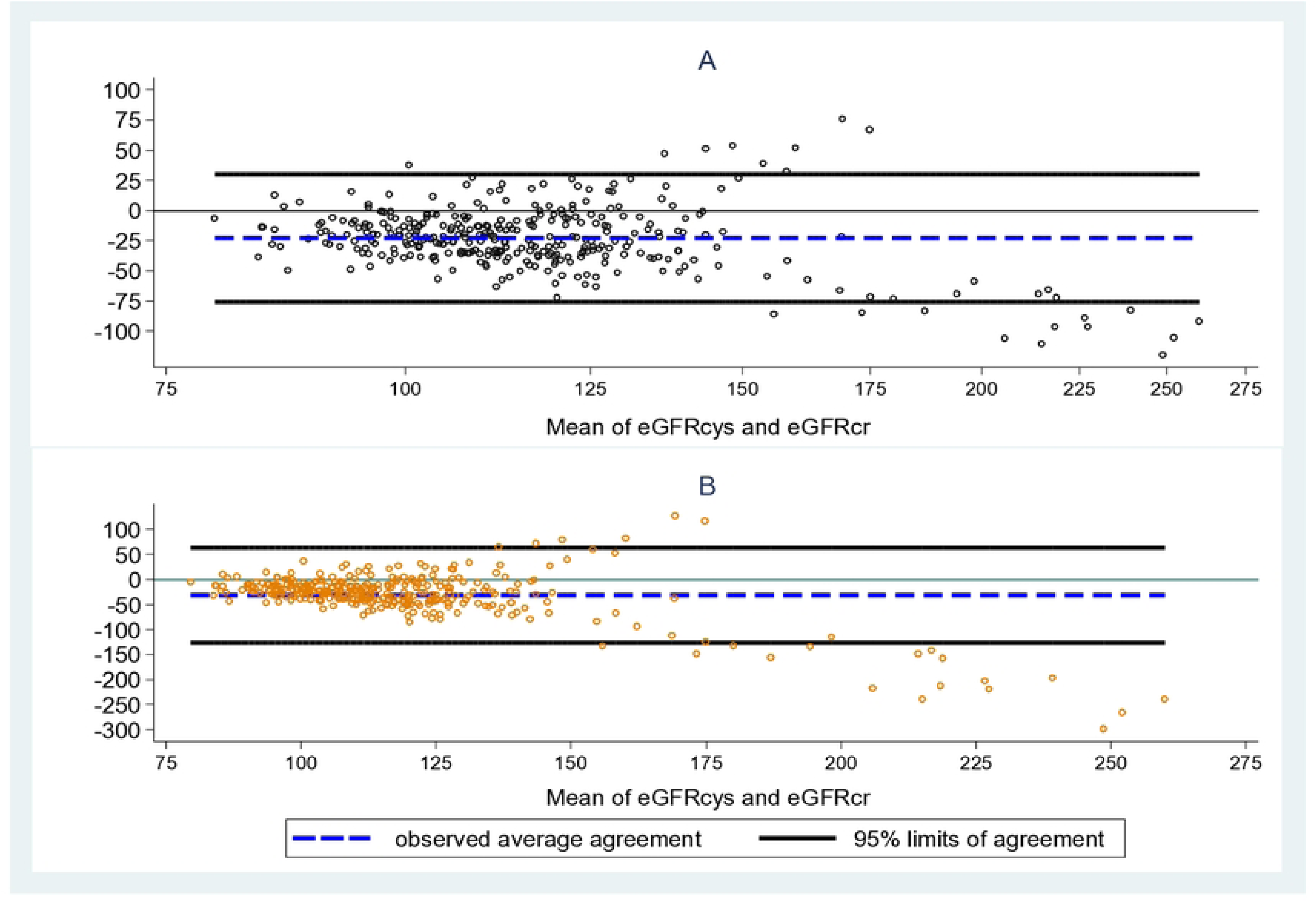
Panel A: Bland-Altman plot featuring percentage differences between eGFR estimated with cystatin C (Berg 2014) and creatinine (Hoste 2014) based formulas. Panel B: Bland-Altman plots showing the agreement between eGFR estimated with cystatin C (Berg 2014) and Creatinine (Hoste 2014) based for1nulas assu1ning horizontal 95% LOA (black lines) and mean bias (blue dash).

For children with both eGFRcys and e GFRcr below 120 ml/min/1.73m², the mean bias percentage for this group was-15%, with LOA ranging from-43.6% to 13.5% (Figure 3-A). The bias between the two methods was-14.8 ml/min/1.73m², and the LOA ranged from-42.6 to 13.1 ml/min/1.73m² (Figure 3-B).

**Figure 3.**
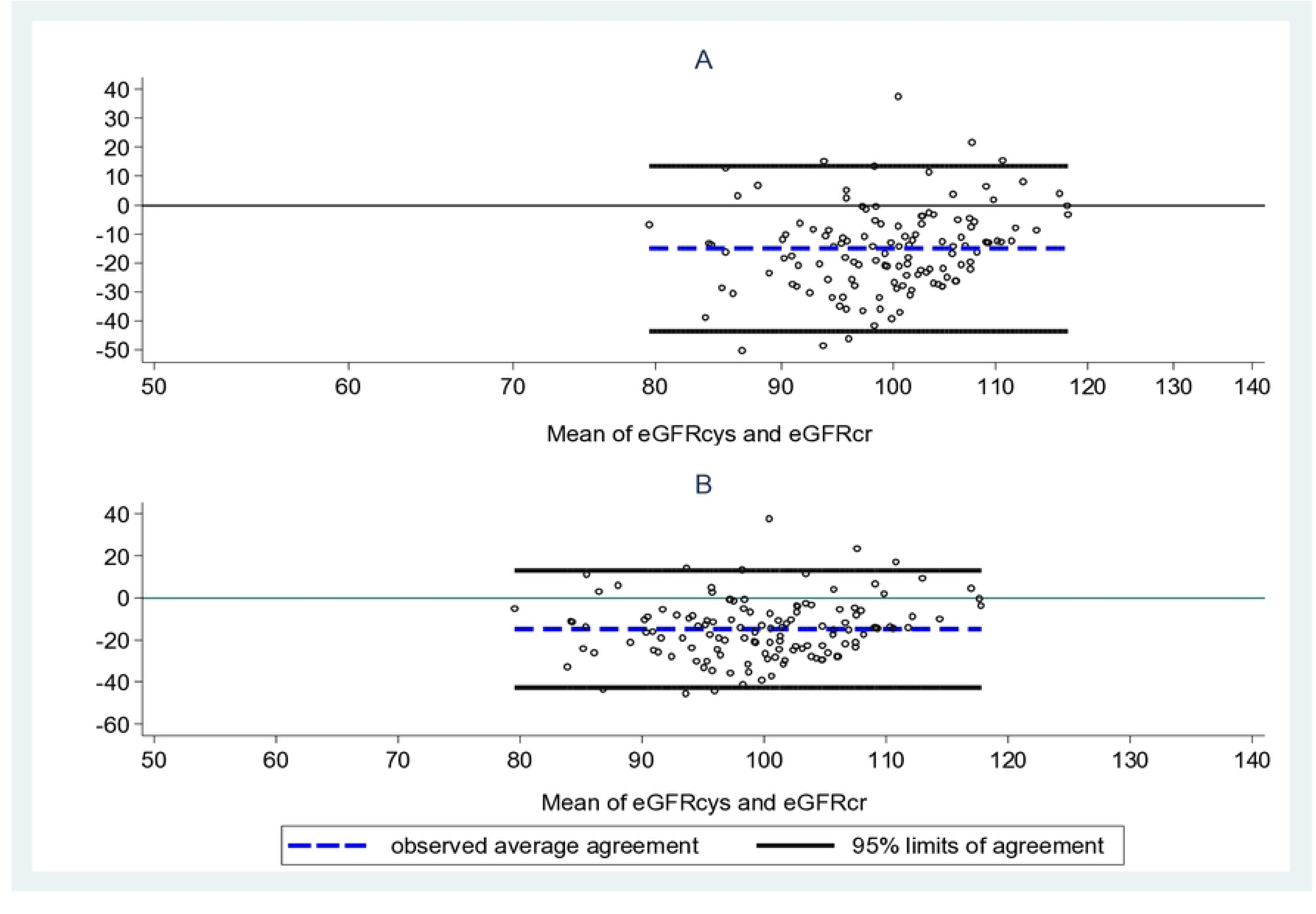
Panel A: Bland-Altman plot of percentage differences in eGFR estimates using cystatin C (Berg 2014) and creatinine (Hoste 2014) formulas for children with eGFR < 120 ml/min/l.73m^2^ Panel B: Bland-Altman plots showing the agreement between eGFR estimated with cystatin C (Berg 2014) and Creatinine (Hoste 2014) based for1nulas for children with eGFR < 120 ml/min/l.73m^2^ assu1ning horizontal 95% LOA (black lines) and mean bias (blue dash).

A sensitivity analysis to investigate the agreement of GFR estimated using formulas used in the current study and locally used creatinine-based Schwartz 2009 formula revealed poor agreement with cystatin C-based Berg 2015 formula (Figure 4-A), but good agreement with eGFR estimated with creatinine-based Hoste 2014 (Figure 4-B). For children with eGFR < 120 ml/min/1.73m², the average bias between Schwartz 2009 and cystatin C-based Berg 2015 formula was-16.2 ml/min/1.73m², with LOA ranged from-44.0 and 11.6 ml/min/1.73m² (Figure 4-C). In contrast, an average bias of-2.6 ml/min/1.73m² with LOA from-12.7 to 7.5 ml/min/1.73m² was observed between creatinine-based Schwartz 2009 and Hoste 2014 formulas (Figure 4-D).

**Figure 4.**
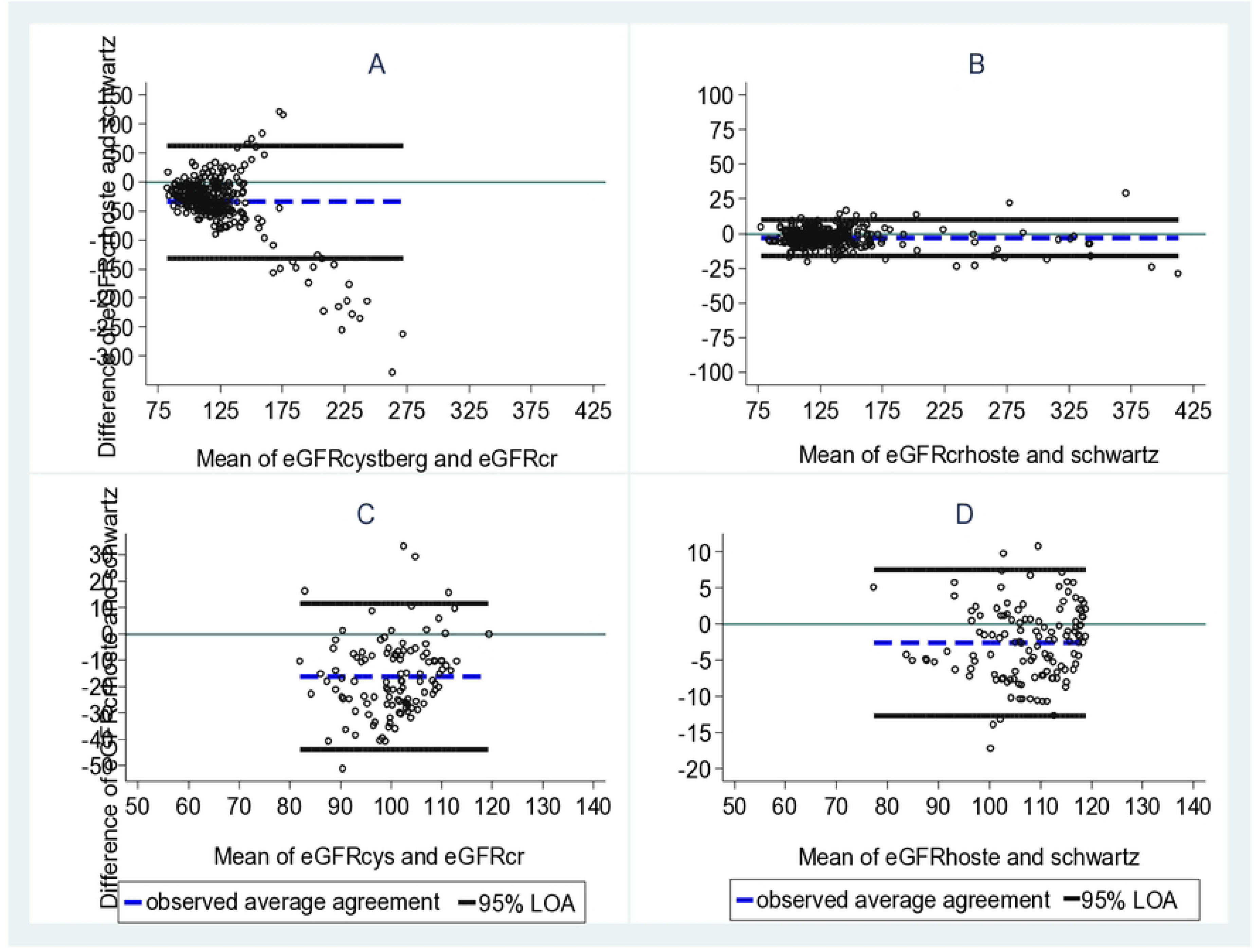
panel A: Bland-Altman plots showing the agreement between eGFR estimated with cystatin C (Berg 2014) and Creatinine based (Schwa1tz 2009) fo1mulas for the whole sample. Panel B: Bland-Altman plots showing the agreement between eGFR estimated with both creatinine based Hoste 2014 and Schwartz 2009 formulas for the whole sa1nple, Panel C: Bland-Altman plots showing the agreement between eGFR estimated with cystatin C (Berg 2014) and Creatinine (Schwartz 2009) based formulas for children with eGFR < 120 ml/min/1.73m^2^. Panel D: Bland-Alttnan plots showing the agreement between eGFR estimated with both creatinine based Hoste 2014 and Schwartz 2009 fo1mulas for children with eGFR < 120 ml/min/l.73m^2^, assuming horizontal 95% LOA (black lines) and mean bias (blue dash).

In the overall cohort, 220 (64.7%) participants had higher eGFRcr compared with eGFRcys (negative eGFRdiff group), 94 (27.6%) had concordant eGFR and 26 (7.7 %) had higher eGFRcys compared with eGFRcr (positive eGFRdiff group). Participants with higher eGFRcr were slightly older and had a higher fat-free mass whereas participants in the higher eGFRcys (positive eGFRdiff) group were younger and a lower fat-free mass (**Table 4).**

**Table 4:** Comparison of characteristics of children at 10-year visit by different level of agreement between cystatin C and creatinine based eGFR (eGFRdiff (eGFRcys-eGFRcr))

| Characteristics | eGFRdiff Group (eGFRcys-eGFRcr) |  |  | P-value |
| --- | --- | --- | --- | --- |
|  | Higher eGFRcr<br>(eGFRdiff < -15<br>mL/min/1.73 m<sup>2</sup>)<br>(N=220) | Concordant<br>(eGFRdiff -15<br>to < 15<br>mL/min/1.73<br>m<sup>2</sup>) (N=94) | Higher eGFRcys<br>(eGFRdiff ≥ 15<br>mL/min/1.73m<sup>2</sup>)<br>(N=26) |  |
| Age (yrs) | 9.9 (1.0) | 9.6 (0.9) | 9.4 (0.8) | 0.01 |
| Sex |  |  |  | 0.45 |
| Male | 118 (53.6) | 44 (46.8) | 15 (57.7) |  |
| Female | 102 (46.4) | 50 (53.2) | 11 (42.3) |  |
| Height (cm) | 132.7 (7.9) | 131.6 (7.2) | 130.3 (6.9) | 0.12 |
| Weight (kg) | 27.6 (5.5) | 26.9 (7.3) | 25.9 (4.4) | 0.29 |
| C-reactive protein | 1.05 (2.8) | 1.3 (4.1) | 1.4 (3.3) | 1.0 |
| Fat mass (kg) | 5.7 (3.2) | 5.4 (4.6) | 5.4 (2.1) | 1.0 |
| Fat-free-mass (kg) | 21.9 (3.3) | 21.6 (3.5) | 20.5 (2.8) | 0.06 |

Data are presented as mean (SD) or n (%).
P-value for positive eGFRdiff versus negative eGFRdiff.

### Factors associated with higher cystatin C and creatinine estimates

After multivariable adjustment, only fat-free mass of the anthropometric, body composition and clinical (C-reactive protein) characteristics at 10-year follow-up exhibited a marginally significant association (P=0.07) with higher eGFRcys (Table 5). A 1 kg increase in fat-free mass decreased the odds of higher eGFRcys by 22%, relative to concordant eGFR, while holding all other variables in the model constant.

**Table 5:** Multinomial regression analysis identifying factors associated with level of agreement between cystain C and creatinine based estimated GFR among children attending the 10-year follow-up.

| Exposures | Level of agreement (eGFRdiff Group (eGFRcys-eGFRcr)) |  |  |  |
| --- | --- | --- | --- | --- |
|  | Higher eGFRcr vs Concordant Category |  | Higher eGFRcys vs Concordant category |  |
|  | Model-1 | Model-2 | Model-1 | Model-2 |
|  | RRR (95% CI) | RRR (95% CI) | RRR (95% CI) | RRR (95% CI) |
| Age (yrs) | 1.44 (1.11; 1.86) | 1.57 (1.12; 2.21) | 0.77 (0.49; 1.21) | 0.83 (0.45; 1.54) |
| Sex (female) | 0.76 (0.47; 1.24) | 0.71 (0.43; 1.18) | 0.63 (0.26; 1.52) | 0.52 (0.21; 1.29) |
| Height (cm) |  | 1.0 (0.92; 1.05) |  | 1.04 (0.92; 1.18) |
| Fat mass |  | 1.03 (0.95; 0.13) |  | 1.08 (0.93; 1.27) |
| Fat-free-mass |  | 0.96 (0.85; 1.09) |  | 0.78 (0.60; 1.02) |
| C-reactive protein |  | 0.97 (0.90; 1.04) |  | 1.0 (0.89; 1.10) |

We visualized the higher eGFRcys group in a scatter plot (Supplemental Figure 2) which showed that these were all at eGFR above 100ml/min. Supplemental analyses suggested that the borderline association with fat-free mass was confounded by eGFR level (Supplemental Tables 3 & 4).

## Discussion

Our findings revealed significant heterogeneity in estimated eGFR distributions depending on the cystatin C or creatinine-based estimation formula used. Even for formulas that appear to be more closely aligned, there is a substantial discrepancy in their estimations. Overall, we observed a poor agreement between eGFRcys and eGFRcr.

The bias increased with rising average eGFR, and it tended to become more negative as GFR increased. The calculated average percentage difference between the two eGFR estimates across the wide range of possible eGFR values was 23.8%; in 80% of cases, this difference was less than 40%. A study has reported that, in cases when the difference between the two measurements is below 40%, a high P30 accuracy rate of more than 90% was documented (7,35). Cystatin C is a more accurate but costly biomarker for pediatric kidney function assessment in high-income settings (13,14,36,37) (14). If it holds true that cystatin C based eGFR is more accurate than creatinine based eGFR, then the creatinine based eGFR may be overestimating kidney function in Ethiopian children on average by 30 ml/min/1.73m^2^. In this study, 64.7% had at least 15 mL/min/1.73 m^2^ lower eGFRcys than eGFRcr. Results among adults from Malawi, Uganda, and South Africa support our findings (21).

In the present study, eGFRdiff was higher than among adults in North America and Europe, where eGFRcys and eGFRcr values differed by more than 15 mL/min/ 1.73 m^2^ in around one-third of participants (11,15,20). A large difference between eGFRcys and eGFRcr indicates a large error compared with mGFR in eGFRcr, eGFRcys, or both. This might be because of clinical conditions that affect the level of creatinine or cystatin C independent of GFR (known as non-GFR determinants) and that, on average, differ from conditions that were present in participants in the studies in which the GFR estimating equations were developed (10). Prior studies reported that individuals who have large negative eGFRdiff (with eGFRcys lower than eGFRcr) might be at higher risk for multiple adverse health outcomes. It will be important to determine whether negative eGFRdiff in children may similarly be associated with higher risk for adverse health outcomes as observed in adults (19,38–40).

The present study revealed a marginally significant negative association between fat-free mass and higher eGFRcys when compared to children in whom eGFRcys and eGFRcr were more comparable (+/-15ml/min/1.73 m^2^ range). We speculate that this association may be an artifact of extremely high eGFRcys values, in which estimation formulas were not developed to detect reliable result in that range and may be less reliable. Hence, the observed poor association likely can be attributed to a range where both formulas are likely not valid, so may not hold great clinical importance.

### Strength and limitations of the study

The strength of this study is that is new information about markers of kidney function among children in a low-income country. Additionally, fat mass and fat-free mass was measured using the accurate and feasible ADP method. However, there are some limitations of our study. First, previous studies have found potential associations of cystatin C with puberty (41), nevertheless we did not have data on puberty, although most children were likely pre-pubertal. Second, we did not use a gold standard GFR measurement to validate the estimation formulas in our study populations due to invasiveness, high cost, and time-consuming nature of the procedure, as well as ethical considerations. Thirdly, the study was done in apparently healthy children from an urban setting who may not be representative of all children in Ethiopia.

In conclusion, this study identified poor agreement between cystatin C and creatinine-based eGFR particularly at high eGFR level. Additionally, the study showed that discordant eGFRcr and eGFRcys was common, with eGFRcys lower than eGFRcr being the typical pattern. The study implies that the routinely utilized creatinine-based GFR estimation formula might over-estimate kidney function among Ethiopian children. A validation study using Inulin urinary clearance, the gold standard for GFR measurement, would be beneficial (42). However, due to its invasive nature and financial concerns in healthy children(6), iohexol plasma clearance studies are recommended as a more practical alternative (43).

## Author contributions

The authors’ responsibilities were as follows– HF, JCKW, TG, RW, DN, SF, DY, and BZ: designed the study; BZ, RA, and BSM: supervised the data collection; HF, JCKW, TG, RW, DY, SF, DN, MFO and MA: participated in methodology; BZ and DN: did the data analysis; BZ: wrote the first draft of the manuscript and had responsibility for the whole work; BZ, DN and DY: had primary responsibility for the final content; All authors contributed to the manuscript revisions and all authors: read and approved the final manuscript.

## Data Availability Statements

The data underlying this article will be shared on reasonable request to the corresponding author.

## Data Availability

The data supporting the findings of this study cannot be made publicly available because the ethical approvals obtained from the Institutional Review Boards of Jimma University and the London School of Hygiene & Tropical Medicine did not include provisions for public data sharing. However, the data may be made available upon reasonable request to the corresponding author.

